# Associations between out of home food sector outlet menu healthiness scores, menu characteristics and energy consumed by customers in 2021-2022

**DOI:** 10.1101/2024.10.24.24316054

**Authors:** Amy Finlay, Yuru Huang, Jean Adams, Andrew Jones, Rebecca Evans, Eric Robinsona

## Abstract

Greater consumption of food prepared outside of the home (OOH) is associated with higher energy intake. Strategies are needed to make eating OOH food less harmful to health. Identifying menu characteristics that contribute to higher energy consumption OOH could aid characterisation of OOH outlets by their relative healthiness and inform future policy intervention in the OOH food sector.

Customers (N=3718) were asked to recall their food orders upon exiting a range of OOH outlets across four local authorities in England during 2021 and 2022. For each outlet, universal health rating scores were calculated based on select menu characteristics and deep learning healthiness scores were calculated based on outlet name. Random forest models and robust linear regression models clustered by outlet were used to identify whether outlet healthiness scores and individual menu characteristics were associated with kcal consumed.

Universal health rating scores, but not deep learning scores, were predictive of energy consumed during OOH outlet visits (-28.27; 95% CI -44.76 to -11.77; p=.003). Menu characteristics with the greatest importance for predicting energy consumed were the percent of savoury main menu items over 600kcal and 1345kcal, the number of desserts, the number of unique vegetables, and the percent of drinks over 100kcal. Menu characteristics accounted for 29% of variance in energy consumed by customers.

Universal health rating scores may be a useful tool to characterise the healthiness of OOH outlets in England. Investigating the potential impact of OOH outlet health ratings on consumer and business behaviour is now warranted.

## 1. Introduction

The consumption of foods and beverages (hereafter: food) prepared outside of the home (OOH) is associated with greater intake of energy, fat, saturated fat, sugar, sodium and protein, and lower intake of micronutrients(Lachat et al., 2012; Powell & Nguyen, 2013). It is therefore unsurprising that UK adults who show increased exposure to and consumption of OOH food typically have a higher BMI and body fat percentage(Albalawi et al., 2022; Burgoine et al., 2014). Individuals who consume OOH food at least once a week are shown to have a greater mean daily energy intake, with both adults and children consuming an additional 55-168kcals compared to those eating OOH food less frequently (Goffe et al., 2017). It is particularly concerning that children from lower socioeconomic backgrounds exhibit greater intake of energy and sugar-sweetened beverages (SSBs) from OOH food outlets(Goffe et al., 2017; Powell & Nguyen, 2013). The consumption of OOH food is therefore a potential contributor to the SEP obesity-related health inequalities.

Strategies are needed to make eating OOH food less harmful to health(Dimbleby, 2021; Obesity Health Alliance, 2021). It is likely that the characteristics of OOH food menus nudge consumers toward unhealthy choices, for example through the large number of unhealthy food options, or the presence of price promotions and meal deals(Dunn et al., 2020; Robinson et al., 2018). Identifying outlet menu characteristics that contribute to higher energy consumption during OOH visits therefore has potential to both characterise the relative healthiness of different OOH outlets and inform interventions in the OOH sector.

It has been suggested that a universal health rating for OOH food outlet menus could help consumers to distinguish between healthier and less healthy outlets, even if they appear to sell very similar products(Goffe et al., 2020). This may be useful when ordering through food delivery platforms as a health rating could inform the outlet choice alongside the already provided hygiene ratings, customer reviews and marketing(Goffe et al., 2020; Riaz et al., 2022). Goffe et al.(Goffe et al., 2020), recruited expert academic researchers in public health and nutrition to rate a range of takeaway outlets according to their healthiness. They then identified menu characteristics that were statistically associated with expert scores. Identified characteristics of food menus associated with (un)healthiness included the number of dessert items, the number of mentions of salad items, the number of mentions of chips, fries or wedges, and the number of price promotions/meal deals(Goffe et al., 2020). Based on Generalised Linear Model fitted scores derived from menu characteristics, cut offs were defined and all outlets given a health rating between 0 and 5.

Informed by the work of Goffe et al.(Goffe et al., 2020), Huang et al.(Huang et al., 2024) modified the rating system and created a deep learning model that could be applied to a wider range of food outlets in the out-of-home food sector (i.e. restaurants, cafes and coffee shops, pubs), specifically those without an online presence where menus were not widely available for analysis. This deep learning model can be used to predict outlet healthiness based on the outlet name alone with absolute difference between predicted and universal health rating values relatively small. Neither the modified universal health rating nor the deep learning model developed by Huang et al., have been tested in terms of their ability to predict energy consumed during an OOH visit.

In addition to understanding how existing overall outlet healthiness scores relate to energy consumed during OOH visits, examining the independent contributions that individual components of menu healthiness ratings and other menu characteristics have on energy consumed will be informative. Experimental evidence has shown that when a larger proportion of healthy food options are offered, people are more likely to make healthier food choices(Langfield et al., 2023). One study examined participant choice and consumption of supermarket ready meals when the proportion of healthy vs less healthy options was altered(Langfield et al., 2023). When participants were provided with a choice of 70% healthy options (vs. 30% healthy in the control condition), significantly lower energy (-196kcals) was consumed and effects were similar across lower and higher SEP participants. Although the nutritional quality of menu items in the OOH food sector(Robinson et al., 2018) are likely poorer than ready meals sold in supermarkets(Remnant & Adams, 2015). We are aware of no research which has examined how availability of higher vs. lower kcal menu options or other menu characteristics in the OOH sector relate to energy consumed during OOH sector visits. Therefore, the primary aims of this study were:

- To identify whether existing outlet healthiness rating tools predict the amount of energy consumed from that outlet during an OOH food eating occasion.
- To identify whether individual characteristics of food menus are associated with energy consumed from that outlet during an OOH food eating occasion.

Secondary aims:

- To examine whether associations differ according to participant socioeconomic position (SEP, measured by level of education).

## 2. Methods

This study was pre-registered on the open science framework https://osf.io/vx9rb/. Ethical approval was granted by the University of Liverpool’s Ethics Committee (Project ID: 10137) and all participants provided informed verbal consent.

### 2.1. Data source

We made use of data collected as part of a project examining consumer purchasing and consumption in large out of home food sector outlets (having ≥250 employees) during August-December of 2021 and 2022. A total of 6548 participants were recruited across two waves of data collection from 330 outlets (76 unique businesses) across four local authorities in England spanning different quintiles of deprivation. For full study information, including sampling, see (Polden et al., 2024). The eligible outlets were stratified by business type and IMD quintile within each local authority sampled. All outlets were classified as one of the following: Restaurants (N=788), Fast-food and takeaways (N=1572), Cafes and coffee shops (N=1217), Entertainment venues (N=121) and Pubs, bars and inns (N=20).

Area-level deprivation of outlets was assessed using the Index of Multiple Deprivation (IMD) which is a measure calculated with consideration of the surrounding area in terms of factors such as income, employment, education and crime(Ministry of housing communities & local government, 2020). To obtain the IMD quintile of individual outlets, we used IMD calculated at the Lower Super Output Area level.

IMD quintiles calculated at a local authority level was used to characterise the local authorities where data collection took place. Local authorities selected were from the North (Liverpool; IMD1), the Midlands (Dudley; IMD2), the South (Milton Keynes; IMD3, IMD4) and London (Richmond; IMD4, IMD5). The project was conducted in 2021 and 2022 as calorie labelling was introduced as a national policy in April 2022 and change in energy consumed in eligible outlets was examined from 2021 (pre policy) vs. 2022 (post policy). However, there was no change in energy consumed in 2021 vs. 2022(Polden et al., 2024).

As part of this study, upon leaving outlets participants were surveyed and asked to report their food purchases and consumption to researchers. In the original study, any participants with missing data were removed, resulting in a sample of n=6409. For approximately 40% of the purchases recorded, online menus of visited outlets were not available to explore menu characteristics, therefore the final sample for the present study was n=3718 participants.

### 2.2. Measures

#### Participant characteristics

Participant characteristics were self-reported as part of the outlet exit survey. Data collected were participant age, gender, ethnicity and highest level of education qualification achieved.

#### Outlet and other measured characteristics

All purchases were categorised by year (2021 vs 2022), outlet type (Restaurants, Fast-food and takeaway, Cafes and coffee shops, Entertainment venues and Pubs, bars and inns), day of the week (weekday vs weekend), time of day (lunch vs dinner). Outlets were also categorised by their Index of Multiple Deprivation (IMD).

#### Consumption

To calculate the energy consumed, participants were questioned regarding the items they purchased for themselves, and any items that were shared or left uneaten. Reported items were linked to MenuTracker data(Huang et al., 2022) which provided the calorie content (kcal) of meals at the time they were purchased. MenuTracker is a database that scrapes nutritional information from online outlet menus for large UK businesses in the OOH food sector. This data is collected quarterly. Where kcal content was not available on MenuTracker, nutritional information was collected from outlet websites between September-November 2022.

### 2.3. Menu Healthiness Scores

#### The modified universal health rating model scores

Goffe et al(Goffe et al., 2020) developed a model to assign menus with an overall healthiness rating using a generalised linear model (GLM). Healthiness scores characterised by the GLM were based on the number of dessert items, the count of all mentions of salad or related items, the count of all mentions of chips/fries/wedges, the number of unique vegetables mentioned, the number of water options, the number of milk options, the number of multi-size options and how these variables related to nutrition experts’ ratings of overall menu healthiness. For this present study, in line with Huang et al. (Huang et al., 2024) we used a modified universal health rating. We modified the existing universal health rating model created by Goffe et al. (Goffe et al., 2020) according to data available, which prevented inclusion of number of ‘multi-size items’ (not in MenuTracker). To examine the impact of exclusion of this item from the universal health rating score, using the data of Goffe et al., we assessed fitted scores of the original Goffe model with the modified model (minus multi-size options) and found the scores to be highly correlated (r(147) = 0.94, p < .001).

Fitted scores in both the original and modified model ranged from 2.6 (least healthy) to 10.0 (healthiest) (Goffe et al., 2020) but in the present study, fitted healthiness scores from a GLM ranged from -0.07 (least healthy) to 12.69 (healthiest). This difference in range was due to the greater range of observed characteristics in the MenuTracker dataset. For example, in the Goffe dataset based on takeaway outlets, the number mentions of chips ranged from 0-48, whereas for menus obtained through Menutracker for the outlets included in the present study this was 0-118. MenuTracker collects information of outlet menu categories, items, and nutritional information from outlet websites. The menu healthiness scores for each outlet were calculated based on the above characteristics. These scores were calculated for 2021 and 2022 menus separately, to account for any changes to menus over the data collection period.

#### Deep learning scores

Huang et al., developed a deep learning model which is informed by modified universal health rating model scores. The deep learning model was developed to predict the healthiness of outlet menus on a large scale and was trained on a number of variables but found to be most accurate at predicting healthiness when using outlet name alone. Where possible, outlet names included the geographical location (for example ‘Starbucks Myrtle Street’). For a number of outlets, the exact location data was not available, so scores were calculated based on the outlet name and broader location (for example ‘Starbucks Liverpool). The final model provides outlets with a score from 0 (least healthy) to 12 (healthiest). These scores are the same for 2021 and 2022 data.

### 2.4. Additional menu characteristics

Further characteristics (to the above) explored were related to the energy content of menu items. Due to the difficulty in quantifying a meal automatically (e.g. create your own meal with a main item and side dishes or small plates rather than a simple starter, main, side dish) all food items were re-categorised into savoury, sweet, sharers, beverages and condiments. All items were categorised by one researcher and 10% of all items across all menus were checked by a second researcher. See Supplementary Material 1 for further information on menu category definitions and example menu items.

We extracted menu categories and energy content from MenuTracker, in order to characterise the proportions of menu items meeting different public health recommendations for energy content. For food items, the proportion of savoury menu items (excluding sharers) that were over 600kcal were calculated. This level was selected as 600kcal is the recommended intake for lunch and dinner meals provided by Public Health England(Public Health England, 2020). This is also the maximum guideline for starters and side plates in the England calorie reduction strategy. Within this same strategy(Public Health England, 2020), 1345kcal was determined as the maximum guideline for calories per portion. The final threshold for main menu items was 2000kcal whereby any items exceeding this would exceed the recommended daily intake for women(NHS, 2023). Desserts were not included in these calculations as several outlets had no, or minimal dessert options. Sharing plates were excluded from these thresholds to focus on meal items intended for one person. After closer examination of the data, we determined that proportion of items over 1345/2000kcal was more meaningful than presence of any items over 1345/2000kcal due to the large number of menu items meeting these thresholds (this is a minor deviation from our pre-registered protocol: https://osf.io/vx9rb/)For beverages, the proportion of beverages that were over 100kcal were recorded. Therefore, the final variables of interest were percent of drinks over 100kcal, percent of savoury menu items over 600kcal, 1345kcal and 2000kcal. Mentions of water were not examined as an individual outlet menu characteristic, as there was only a small number of outlets with any mentions of water.

### 2.5. Analysis

Robust linear regression models (x2) clustered by outlet were used to examine associations between each of the healthiness scores and energy consumed. Age, gender (male vs female), ethnicity (white vs other), and SEP (High (degree and above) vs Low) were included in demographic adjusted models. Outlet type (café, restaurant, pub, fast food, entertainment) and IMD were included as restaurant/location control variables and time (pre/post kcal labelling, i.e. 2021 vs. 2022), time of day (lunch vs dinner) and day (weekday vs weekend) were included in the models. A robust linear regression model clustered by outlet was also used to examine the independent associations of specific menu characteristics (which made up the menu healthiness score) with energy consumed. Details of this model are available in Supplementary Material 2.

#### 2.5.1. Secondary analyses

A random forest model was conducted to examine whether the individual menu characteristics (components of the universal health rating and energy thresholds described above) predicted energy consumed. This model was selected to determine feature importance and identify the most accurate group of menu characteristics for predicting energy consumed. To control for outlet type, all outlet types were dummy coded individually. As all features were deemed important in an initial model (Supplementary Material 3), a robust linear regression model was conducted with all features to assess multicollinearity.

Multicollinearity was detected for four characteristics: percent of items over 1345kcal (VIF 10.66), mentions of chips (VIF 7.76), Fast food outlets (VIF 6.31) and percent of items over 2000kcal (VIF 6.05). Mean centring the variables did not improve multicollinearity, so variables were removed in reverse order of feature importance until all VIFs were an acceptable level. The percent of items over 2000kcal was removed first, followed by mentions of chips, as removing fast food had a negligible impact on the other variables. All remaining variables had a VIF below 5 so were deemed acceptable. The model with collinear variables removed is shown in Supplementary Material **4**.

#### 2.5.2. Exploratory analyses

Moderation analyses were conducted to identify whether the relationship between healthiness scores and energy consumed differed according to outlet type (i.e. restaurant vs entertainment venue) or participant level of education (low vs high), as there is some evidence that energy intake in the OOHF sector differs according to socioeconomic position and type of OOH source (e.g. eating out vs takeaway)(Goffe et al., 2017). One menu characteristic (mentions of chips) was associated with energy consumed (see Results section), so further analyses identified whether this association was moderated by education or outlet type.

Results for primary analyses were considered significant at p<0.05. To account for multiple comparisons results for secondary and exploratory analyses were considered significant at p<0.01. All analyses were conducted in R with packages: performance (Lüdecke et al., 2021), estimatr (Blair et al., 2022) and car (Fox & Weisenberg, 2019) for regression models and MLmetrics (Yachen, 2024) and Boruta (Kursa & Rudnicki, 2010) for random forest models.

## 3. Results

### 3.1. Demographics

N=3718 participants were included in the final sample. Of all participants included, 53% (n=1958) were women, 79% (n=2940) were White and 55% (n=2064) were classified as having attained a lower level of education (A-level equivalent or below). Full participant demographic data is available in Table 1.

**Table 1:** Participant characteristics.

|  | *N* | % |
| --- | --- | --- |
| *Gender* |  |  |
| *Woman* | 1958 | 53% |
| *Man* | 1760 | 47% |
| **Ethnicity** |  |  |
| *White* | 2940 | 79% |
| *Non-White* | 778 | 21% |
| **Education** |  |  |
| *Low (A level and below)* | 2064 | 56% |
| *High* | 1654 | 44% |
|  | **Mean (sd)** | **Range** |
| **Age** | 40.58 (17.48) | 16-87 |

### 3.2. Menu characteristics

The mean deep learning score for outlets where purchases had been made was 6.72 (±1.00) but ranged from 1.81 (least healthy) to 9.52 (healthiest) on a scale of 0 to 12. The mean menu healthiness score based on menu characteristics was 6.32 (±1.83) and ranged from -0.07 (least healthy) to 12.69 (healthiest).

The descriptive statistics for all menu characteristics measured are shown in Table 2.

**Table 2:**
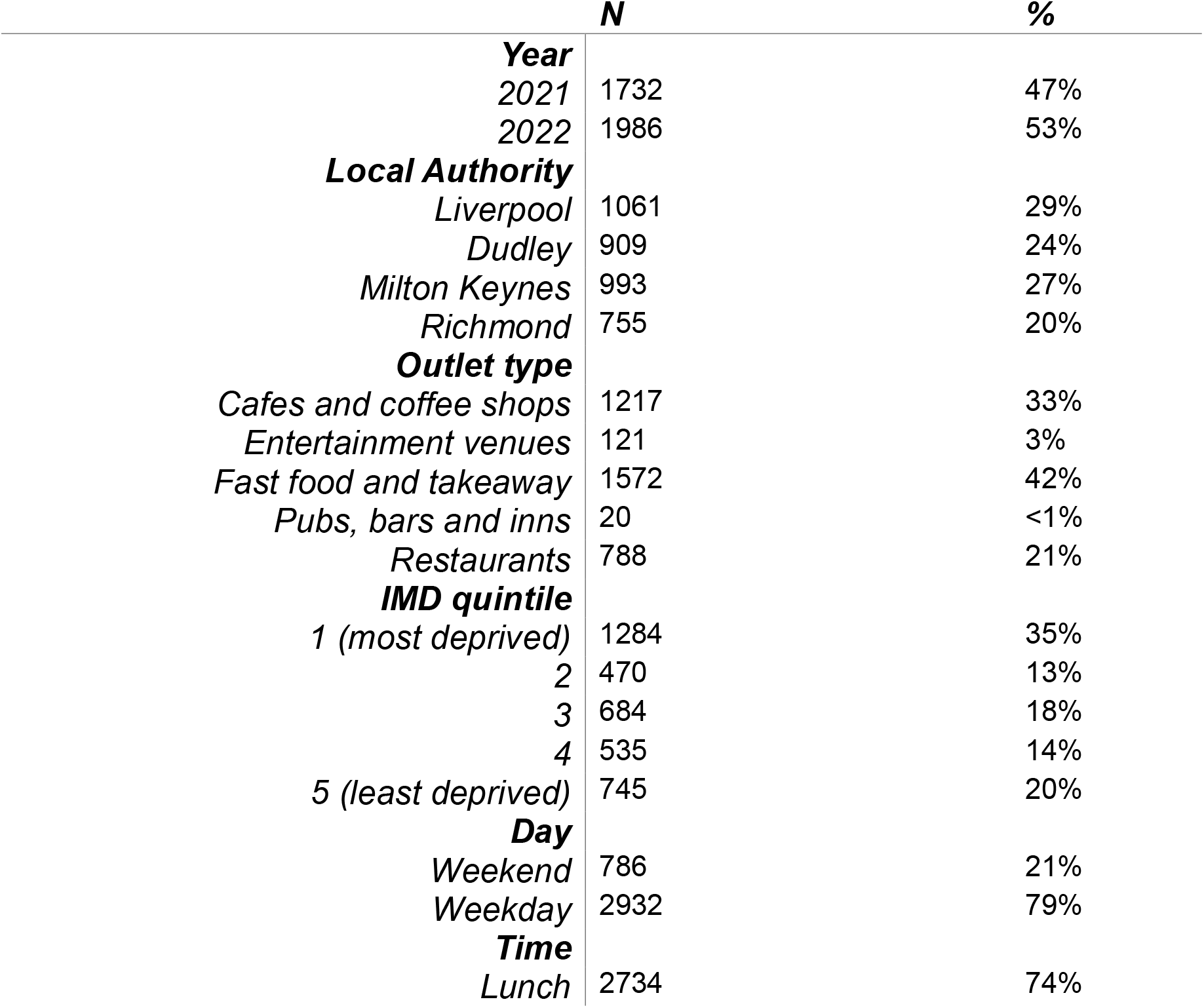
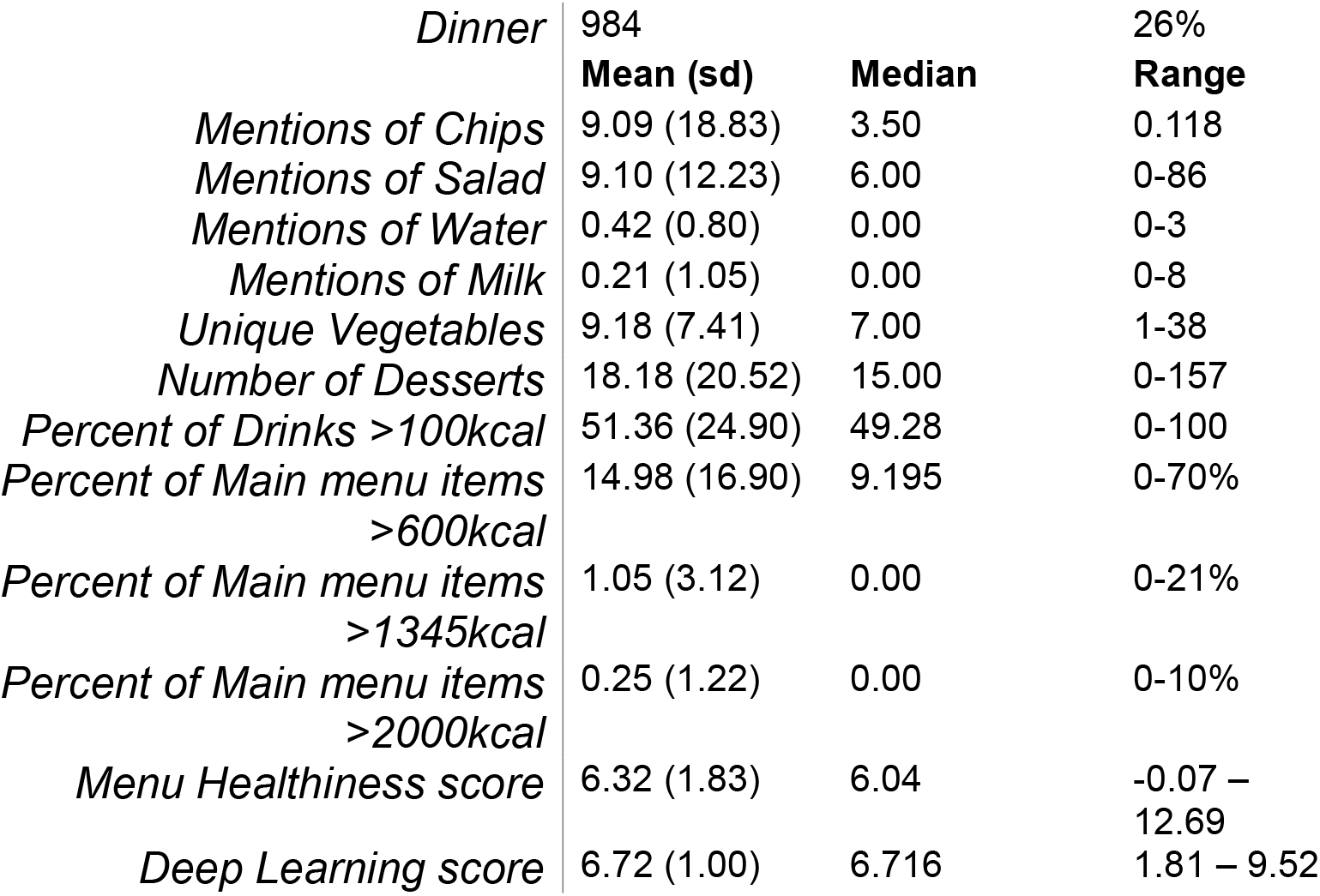
Menu and situational characteristics.

|  | *N* | *%* |
| --- | --- | --- |
| **Year** |  |  |
| *2021* | 1732 | 47% |
| *2022* | 1986 | 53% |
| **Local Authority** |  |  |
| *Liverpool* | 1061 | 29% |
| *Dudley* | 909 | 24% |
| *Milton Keynes* | 993 | 27% |
| *Richmond* | 755 | 20% |
| **Outlet type** |  |  |
| *Cafes and coffee shops* | 1217 | 33% |
| *Entertainment venues* | 121 | 3% |
| *Fast food and takeaway* | 1572 | 42% |
| *Pubs, bars and inns* | 20 | <1% |
| *Restaurants* | 788 | 21% |
| **IMD quintile** |  |  |
| *1 (most deprived)* | 1284 | 35% |
| *2* | 470 | 13% |
| *3* | 684 | 18% |
| *4* | 535 | 14% |
| *5 (least deprived)* | 745 | 20% |
| **Day** |  |  |
| *Weekend* | 786 | 21% |
| *Weekday* | 2932 | 79% |
| **Time** |  |  |
| *Lunch* | 2734 | 74% |

|  |  |  |  |
| --- | --- | --- | --- |
| *Dinner* | 984 |  | 26% |
|  | **Mean (sd)** | **Median** | **Range** |
| *Mentions of Chips* | 9.09 (18.83) | 3.50 | 0-118 |
| *Mentions of Salad* | 9.10 (12.23) | 6.00 | 0-86 |
| *Mentions of Water* | 0.42 (0.80) | 0.00 | 0-3 |
| *Mentions of Milk* | 0.21 (1.05) | 0.00 | 0-8 |
| *Unique Vegetables* | 9.18 (7.41) | 7.00 | 1-38 |
| *Number of Desserts* | 18.18 (20.52) | 15.00 | 0-157 |
| *Percent of Drinks >100kcal* | 51.36 (24.90) | 49.28 | 0-100 |
| *Percent of Main menu items >600kcal* | 14.98 (16.90) | 9.195 | 0-70% |
| *Percent of Main menu items >1345kcal* | 1.05 (3.12) | 0.00 | 0-21% |
| *Percent of Main menu items >2000kcal* | 0.25 (1.22) | 0.00 | 0-10% |
| *Menu Healthiness score* | 6.32 (1.83) | 6.04 | -0.07 – 12.69 |
| *Deep Learning score* | 6.72 (1.00) | 6.716 | 1.81 – 9.52 |

### 3.3. Purchase and consumption

Participants ordered a mean of 864.20 ±553.04 kcals and reported consuming on average

775.90 ±480.06 kcals. As results were the same for both kcals purchased and consumed, only kcal consumption data is reported here.

### 3.4. Associations between outlet healthiness scores and kcal consumed

For both primary models, the VIF of all predictors was below 2, so any influence of multicollinearity was deemed negligible. A robust linear regression clustered by outlet found no association between deep learning scores and kcal consumed (p=.719) (Table 3). However, a significant association was observed between menu healthiness scores and kcal consumed (p<.01) (Table 3).

**Table 3:** Associations between deep learning scores (top), menu healthiness scores (bottom) and kcal consumed.

| *Predictors\** | **Kcal consumed** |  |  |
| --- | --- | --- | --- |
|  | *Estimates* | *CI* | *p* |
| (Intercept) | 677.79 | 432.63 – 922.94 | **<0.001** |
| Deep learning score | -6.00 | -38.66 – 26.66 | 0.719 |
| **Outlet type: Entertainment venues | -76.15 | -189.62 – 37.31 | 0.188 |
| **Outlet type: Fast Food and Takeaways | 196.34 | 128.01 – 264.66 | **<0.001** |
| **Outlet type: Pubs, bars and inns | 689.72 | -18.06 – 1397.49 | 0.056 |
| **Outlet type: Restaurants | 645.46 | 564.76 – 726.16 | **<0.001** |
| ***Outlet IMD quintile 2 | -35.05 | -134.87 – 64.77 | 0.491 |
| ***Outlet IMD quintile 3 | -54.45 | -138.72 – 29.82 | 0.205 |
| ***Outlet IMD quintile 4 | -87.86 | -167.38 - -8.35 | **0.030** |
| ***Outlet IMD quintile 5 | -64.16 | -136.63 – 8.31 | 0.083 |
| Observations | 3718 |  |  |
| R<sup>2</sup> / R<sup>2</sup> adjusted | 0.316 / 0.313 |  |  |
| (Intercept) | 827.416 | 693.84 – 960.47 | **<0.001** |
| Menu healthiness scores | -28.27 | -44.76 – -11.77 | **0.003** |
| **Outlet type: Entertainment venues | -144.96 | -273.12 – -16.80 | **0.027** |
| **Outlet type: Fast Food and Takeaways | 204.80 | 147.08 – 262.52 | **<0.001** |
| **Outlet type: Pubs, bars and inns | 633.93 | -117.16 – 1385.01 | 0.111 |
| **Outlet type: Restaurants | 664.72 | 590.21 – 739.24 | **<0.001** |
| ***Outlet IMD quintile 2 | -33.69 | -133.03 – 65.65 | 0.506 |
| ***Outlet IMD quintile 3 | -50.04 | -130.52 – 30.45 | 0.223 |
| ***Outlet IMD quintile 4 | -94.91 | -173.32 - -16.50 | **0.018** |
| ***Outlet IMD quintile 5 | -72.20 | -137.54 - -6.86 | **0.030** |
| Observations | 3718 |  |  |
| R<sup>2</sup> / R<sup>2</sup> adjusted | 0.326 / 0.323 |  |  |

\*This model controlled for participant characteristics (age, gender, ethnicity, education) and situational characteristics (year, time of day, weekday/weekend).
\*\*Reference value for outlet type is Cafes and coffee shops
\*\*\*IMD = Index of Multiple Deprivation, quintile 1 (most deprived) is the reference value

The interaction between level of education and the two menu healthiness scores was explored in a second step of each of the primary models. Interactions were found to be non-significant for both menu healthiness scores (ps>.750). Full detail of this analysis can be found in Supplementary Material 5).

Interactions were explored between menu healthiness scores and outlet type. A significant interaction was observed between deep learning scores and outlet type. Exploration of both scores according to outlet types individually are shown in Supplementary Material 6. In short, no significant associations were observed between deep learning scores and kcal consumed across the different outlet types (ps>.073). However, for menu healthiness scores, significant negative associations were observed for fast food outlets (-188.71; 95% CI -247.11 to - 130.31; p<.001) and for restaurants (-29.47; 95% CI -51.60 to -7.34; p<.01), but not cafés, pubs, and entertainment venues. Whereby as healthiness scores increased (outlets were scored as healthier), kcal consumed decreased and this effect was greater for fast food outlets than for restaurants.

A further robust clustered linear regression model was conducted to see which of the menu characteristics that make up menu healthiness scores were associated with energy consumed. This model included the 6 menu characteristics as predictors alongside the same covariates included in prior primary analyses. The number of mentions of chips was significantly associated with kcal consumed. Full details are in Supplementary Material 2.

### 3.5. Identifying important predictors

A random forest model identified that all studied features were important predictors of kcal consumption (See Figure 1). A number of collinear predictors were identified, and results were similar with these variables excluded (see Supplementary Material 4). Features with the greatest importance, and therefore contributed most to the accuracy of the model were: the percent of main menu items over 600kcal and 1345kcal, menu items from restaurants (as opposed to cafes and coffee shops, fast food outlets, entertainment venues and pubs), the percent of drinks over 100kcal and the number of desserts and unique vegetables mentioned on menus. Full details of feature importance are available in Supplementary Material 3.

**Figure 1:**
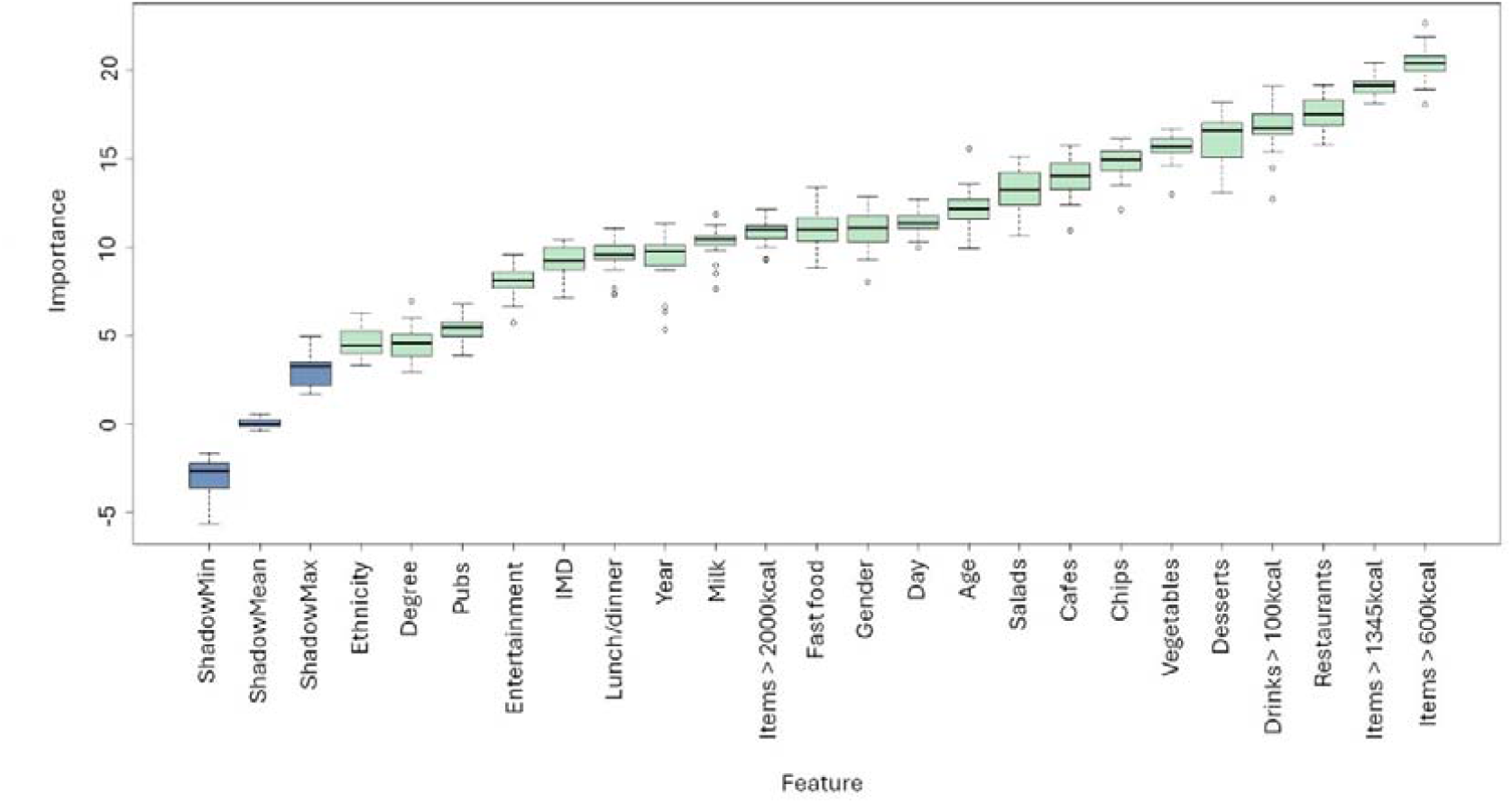
Random Forest Model feature importance with all predictors.

A two-step regression with all characteristics deemed important in the absence of multi-collinearity identified that menu characteristics alone explained 28.9% of variance (Supplementary Material 7), and when participant (e.g., gender) and situational characteristics (e.g., time of day) were included in the model, this increased to 38.8%. Table 4 shows the final regression model whereby the number of desserts, the proportion of main menu items over 600kcal and 1345kcal and the proportion of drinks over 100kcal were positively associated with kcal consumed. The number of mentions of salad were negatively associated with kcal consumed.

**Table 4:**
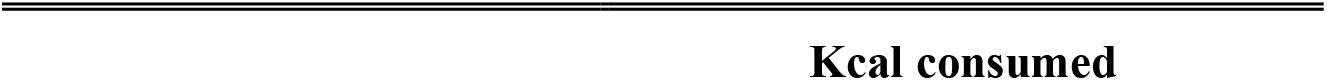
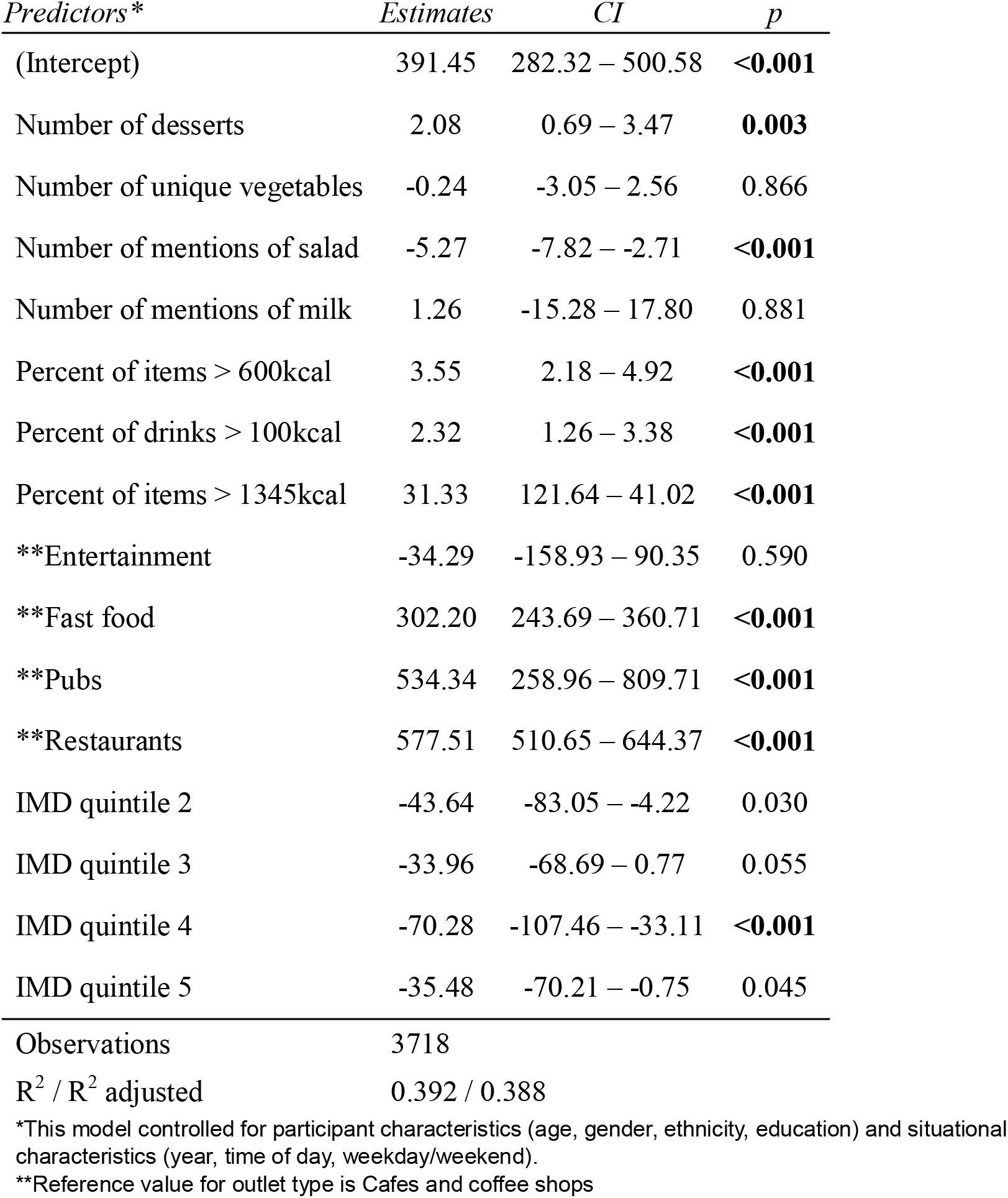
Final regression model with menu characteristics and participant/situational characteristics.

| *Predictors\** | *Estimates* | *CI* | *p* |
| --- | --- | --- | --- |
| (Intercept) | 391.45 | 282.32 – 500.58 | **<0.001** |
| Number of desserts | 2.08 | 0.69 – 3.47 | **0.003** |
| Number of unique vegetables | -0.24 | -3.05 – 2.56 | 0.866 |
| Number of mentions of salad | -5.27 | -7.82 – -2.71 | **<0.001** |
| Number of mentions of milk | 1.26 | -15.28 – 17.80 | 0.881 |
| Percent of items > 600kcal | 3.55 | 2.18 – 4.92 | **<0.001** |
| Percent of drinks > 100kcal | 2.32 | 1.26 – 3.38 | **<0.001** |
| Percent of items > 1345kcal | 31.33 | 121.64 – 41.02 | **<0.001** |
| **Entertainment | -34.29 | -158.93 – 90.35 | 0.590 |
| **Fast food | 302.20 | 243.69 – 360.71 | **<0.001** |
| **Pubs | 534.34 | 258.96 – 809.71 | **<0.001** |
| **Restaurants | 577.51 | 510.65 – 644.37 | **<0.001** |
| IMD quintile 2 | -43.64 | -83.05 – -4.22 | 0.030 |
| IMD quintile 3 | -33.96 | -68.69 – 0.77 | 0.055 |
| IMD quintile 4 | -70.28 | -107.46 – -33.11 | **<0.001** |
| IMD quintile 5 | -35.48 | -70.21 – -0.75 | 0.045 |
| Observations | 3718 |  |  |
| R<sup>2</sup> / R<sup>2</sup> adjusted | 0.392 / 0.388 |  |  |

\*This model controlled for participant characteristics (age, gender, ethnicity, education) and situational characteristics (year, time of day, weekday/weekend).
\*\*Reference value for outlet type is Cafes and coffee shops

## 4. Discussion

This study explored whether menu healthiness scores and menu characteristics of OOH food outlets in England were associated with energy consumed by customers in outlets belonging to large businesses. Menu healthiness scores which were based on the presence of outlet menu characteristics were negatively associated with energy consumed. Outlet menu healthiness scores derived only from outlet name and location using a deep learning model (used to predict menu healthiness scores) were not significantly associated with energy consumed. A range of specific menu characteristics (number of desserts, number of unique vegetables, number of mentions of salad, milk, percent of menu items over 600kcal and 1345kcal and percent of drinks over 100kcal) accounted for 29% of the variation in energy consumed from OOH outlets.

A scoring system for out of home food outlets was developed in previous research (Goffe et al., 2020; Huang et al., 2024) in the context of supporting consumers to make healthier choices in the OOH food sector, and when ordering food from online settings. In the present study, the menu healthiness scoring method adapted from Goffe(Goffe et al., 2020) was a significant predictor of kcal consumption, particularly for fast food outlets and restaurants. This scoring method was initially developed using fast food and takeaway outlet menus and the significant association with kcal consumed in these outlets in particular may be a result of using the measure as it was created, as opposed to pubs, cafes and coffee shops, and entertainment venues in which we did not find healthiness scores were associated with customers’ energy consumption. Conversely, deep learning model scores were not a strong predictor of kcal consumption in this study overall, or for individual outlet types. It is likely that the data used in this model (i.e. outlet name and location) were not sufficient to capture the nuances in food offerings across individual outlets, and across different data collection periods, as the menu healthiness scores were able to. This model was developed with the aim of ranking outlets in terms of their healthiness to identify areas where more unhealthy outlets were present(Huang et al., 2024). It is possible that this method of scoring would have a greater association with energy consumed when examining ranking of outlets rather than absolute scores of outlets. Further work could explore how healthiness can be determined in outlets not classed as fast food or takeaway to more accurately categorise and guide a wider range of outlet types.

Menu characteristics with the greatest importance for predicting kcal consumption were the percent of main menu items over 600kcal and 1345kcal, the percent of drinks over 100kcal, the number of desserts and the number of unique vegetables. The cut-offs selected for main menu items were based on existing UK government guidelines for OOH food. Specifically, 600kcal is the recommended intake for lunch and dinner meals and the maximum guideline for starters and side plates in the UK government’s calorie reduction strategy(Public Health England, 2020). 1345kcal is the maximum guideline for calories per portion according to Public Health England (Public Health England, 2020). Primary analysis of menu characteristics identified that the number of mentions of chips on an outlet menu was positively associated with energy consumed from that outlet. This is perhaps expected as chips are a typically unhealthy item, and a frequent accompaniment in the OOH food sector.

Focusing on the key predictive menu features could guide businesses in the OOH food sector in offering healthier choices. For example, through reformulation or reducing the proportion of main menu items over 600kcal and 1345kcal and drinks over 100kcal while increasing the number of unique vegetables on the menu may be effective strategies to reduce overall energy consumption. This is supported by research showing that a greater proportion of healthier items on a menu lead to lower energy consumption, for individuals of high and low SEP(Langfield et al., 2022). Alternatively, Goffe et al. (Goffe et al., 2020) argue that including a healthiness score on online food delivery platforms per outlet could act as an intervention for consumers, as scores could prompt avoidance of outlets with extremely unhealthy scores. Equally, if outlets were presented on online platforms in order of their healthiness score, this could lead to a greater likelihood of healthier outlet choice. For outlets, providing this type of information would have minimal costs. However, even among outlets with higher healthiness scores, a relatively large number of menu items will typically be of low nutritional quality. Furthermore, the potential for a positive impact of such an intervention still relies on individuals being motivated by health when considering which outlets to order from, although this would be lessened if a scoring system led to structural changes to online platforms (i.e. through re-ordering of outlets). Research suggests that consumers are often not motivated by health when making food-related decisions, particularly if from a lower SEP background(Robinson et al., 2022) or if purchasing OOH food as a ‘treat’(Miura & Turrell, 2014). Therefore, interventions on the healthiness of menus would likely have a greater impact if focused on changing business behaviour rather than consumer choice.

Exploration of whether outlet healthiness scores impact food outlet choice by consumers, specifically for fast food and takeaway outlets may now be warranted. Exploring whether such scores also have an upstream effect on business behaviour will also be vital to assessing potential benefits of this approach. Further work could guide improvements in the healthiness of the OOH food sector through developing guidelines for outlet menus. Nutritional standards exist in certain settings such as schools and hospitals to ensure that nutritious food is served(Department for Education, 2023; NHS England, 2022), however implementing such standards in non-government mandated settings would likely be more challenging. At present it may be more plausible to improve the healthiness of outlets in the OOH food sector by local level policies. For example, the Recipe 4 Health scheme in Lancashire County Council(Lancashire County Council, n.d.) awards outlets that meet achievable standards relating to healthy eating and sustainability.

### 4.1. Strengths and Limitations

This study is the first to investigate whether existing overall menu healthiness scoring methods and individual characteristics are associated with energy consumed in the OOH food sector. A key strength of this study is that MenuTracker data (the data used to determine the energy content of meals) is collected quarterly. This means that the kcal purchased/consumed and outlet menu characteristics are likely accurate to the time of measurement. Additionally, this study used real purchase data and so purchases reflect real world conditions, not experimental ones. A number of the additional menu characteristics measured (%>600, 1345, 2000) measured savoury items only, as there were a number of outlets with no or a very small number of desserts. It is possible that inclusion of these categories could have altered findings. Similarly, due to the large number of outlets with no mentions of water, this variable was excluded from analyses, despite being shown by Goffe et al to be a significant predictor of energy.

In calculating deep learning scores, the exact geographical location could not be obtained for all outlets. This meant that names such as ‘Starbucks Liverpool’ was used rather than ‘Starbucks Myrtle Street’. While this is expected to have limited impact on the findings, it is possible that the deep learning model may be more effective with exact pinpointing of outlet location, or equally without location data at all. Finally, the findings here relate to food purchases in large food chains that were subject to the calorie labelling policy, therefore our findings are not representative of all OOH outlets in the UK. Evidence suggests that meals from independent takeaway businesses in the UK are highly calorific with poor nutrient composition(Jaworowska et al., 2014). If the overwhelming majority of smaller out of home business outlets offer predominantly unhealthy food, then menu healthiness ratings may have limited utility in predicting energy consumed by customers. As such outlets make up a large proportion of the OOH food sector in England this is an important avenue for future research.

## 5. Conclusions

Universal health rating scores of OOH food outlets are likely a useful tool to predict energy consumed in OOH food settings in England. Investigating the potential impact of OOH outlet health ratings on consumer and business behaviour is now warranted. A number of outlet menu characteristics were found to be associated with energy consumption and OOH food sector policies which address these characteristics may benefit public health.

## Data Availability

Data will be made available on the Open Science Framework upon publication

