## Supplementary Material for "Associations between out of home food sector outlet menu healthiness scores, menu characteristics and energy consumed by customers in 2021-2022"

**Supplementary Material 1: Menu item categorisation.**

The MenuTracker tool scrapes information from online outlet menus. Part of the information scraped is saved as the ‘menu section’. While for a minority of outlets the menu categories were clear (e.g. starter, side, main, meat & fish, pizza, pasta, dessert), most outlets had inconsistent groups that were ambiguous and needed further clarification (e.g. bar bites, brunch, bowls) or spanned multiple food categories (e.g. Sunday lunch, gluten free, specials).

To make clearer categories for comparisons across outlets, all items were grouped into sweet, savoury, drinks, condiments, sharers. One researcher worked through all provided menu sections and where necessary individual items to group items into the 5 categories with the following definitions and examples:

- Sweet: Any sweet item intended to be eaten alone (e.g. as a dessert)
  - Menu section contains ‘dessert’ or ‘sweet’
  - Product description indicates a sweet dish (e.g. cake, ice cream, fruit)
- Savoury: Any savoury menu item that could be considered a starter, side, or main dish. Sometimes these items will be small if they are from menus where the customer can design their own meal (e.g. choose your own cooked breakfast could include individual items egg, bacon, tomato, mushroom…)
  - Menu section contains ‘main’, ‘side’, ‘starter’, ‘small plate’, ‘pizza’, ‘pasta’, ‘salad’, ‘meat’, ‘fish’, ‘bread’, ‘burger’, ‘curry’ etc.
  - Product description can be considered a savoury item
- Drinks: Any menu items that could be considered a drink as a standalone item
  - Menu section contains ‘drink’ or ‘beverage’ or specific drink categories e.g. smoothies, milkshakes, coffees
  - Product description is a standalone drink (e.g. americano, berry smoothie, fanta)
- Condiments: Any menu items intended as an addition to a meal item (this does not include food ‘add-ons’ that could be seen as a side (e.g. extra chicken, include fries)
  - Menu section contains ‘sauce’
  - Product description contains ‘sauce’, ‘gravy’, ‘jam’, ‘dressing’, ‘syrup’ etc. as a standalone item
- Sharers: Any menu items intended for more than one person
  - Menu section contains the word ‘sharer’ or ‘sharing’
  - Product description contains the word ‘sharer’ or ‘sharing’ or states ‘for X people’

Two researchers randomly checked 10% of the categorisations and where there were any mis-categorised items, key words were identified that would identify any other discrepancies in the dataset. Once these changes were done, both researchers checked another random 10% of the sample and agreement was perfect.

**Supplementary Material 2: Associations between menu characteristics and kcals consumed**

A robust linear regression model clustered by outlet was used to explore associations between individual components of menu healthiness rating and kcal consumed (Supplementary Table 1). This model included the 6 menu characteristics as predictors alongside the same covariates that were included in primary analyses.

*Supplementary Table 1: Association between menu characteristics that menu healthiness scores are derived from and kcal consumed*

|  | **Kcal consumed** | | |
| --- | --- | --- | --- |
| *Predictors** | *Estimates* | *CI* | *p* |
| (Intercept) | 599.50 | 450.13 – 748.87 | **<0.001** |
| Number of desserts | 3.79 | 0.73 – 6.84 | **0.015** |
| Number of unique vegetables | -2.74 | -9.16 – 3.69 | 0.404 |
| Number of mentions of salad | -3.71 | -12.25 – 4.84 | 0.395 |
| Number of mentions of chips | 0.98 | -2.65 – 4.60 | 0.597 |
| Number of mentions of milk | 14.04 | -18.90 – 46.98 | 0.404 |
| **Outlet type: Entertainment venues | -234.19 | -412.90 – -55.47 | **0.010** |
| **Outlet type: Fast Food and Takeaways | 264.83 | 140.90 – 388.76 | **<.001** |
| **Outlet type: Pubs, bars and inns | 716.40 | -53.64 – 1486.43 | 0.068 |
| **Outlet type: Restaurants | 667.09 | 586.80 – 747.38 | **<0.001** |
| Observations | 3718 | | |
| R^2^ / R^2^ adjusted | 0.335 / 0.331 | | |

*This model controlled for participant characteristics (age, gender, ethnicity, education) and situational characteristics (year, time of day, weekday/weekend, outlet IMD).
**Reference value for outlet type is Cafes and coffee shops

In the above model, multicollinearity was detected for outlet type (VIF=6.41) and chips (VIF=6.17). When outlet type was removed from the model, the VIFs for all other variables were below 4.7 so deemed acceptable. The final model (Supplementary Table 2) indicated that mentions of chips (p<.01) was positively associated with kcal consumed.

*Supplementary Table 2: Association between menu characteristics and kcal consumed (minus collinear predictors)*

|  | **Kcal consumed** | | |
| --- | --- | --- | --- |
| *Predictors** | *Estimates* | *CI* | *p* |
| (Intercept) | 901.95 | 777.98 – 1025.92 | **<0.001** |
| Number of desserts | -1.79 | -5.02 – 1.44 | 0.277 |
| Number of unique vegetables | 5.26 | -1.46 – 11.98 | 0.125 |
| Number of mentions of salads | -0.04 | -7.33 – 7.25 | 0.992 |
| Number of mentions of chips | 6.57 | 1.55 – 11.58 | **0.010** |
| Number of mentions of milk | -8.74 | -43.59 – 26.10 | 0.623 |
| Observations | 3718 | | |
| R^2^ / R^2^ adjusted | 0.161 / 0.158 | | |

*This model controlled for participant characteristics (age, gender, ethnicity, education) and situational characteristics (year, time of day, weekday/weekend, outlet IMD).

No significant interactions between the characteristics deemed significant (mentions of chips) and education were identified (p=.107). A significant interaction was observed restaurants (9.06; 95% CI 3.56 to 14.56; p<.01) but not for entertainment venues, fast food outlets or cafes. Whereby as the number of mentions of chips increased, kcal consumed increased.

**Supplementary Material 3: Final random forest feature importance (all confirmed important)**

|  | Mean Importance | Median Importance | Minimum Importance | Maximum Importance | Norm Hits | Decision |
| --- | --- | --- | --- | --- | --- | --- |
| Day* | 11.89 | 11.78 | 9.88 | 14.34 | 1.00 | Confirmed |
| Lunch/dinner** | 11.61 | 11.55 | 9.99 | 13.69 | 1.00 | Confirmed |
| Year*** | 9.70 | 9.72 | 6.74 | 11.67 | 1.00 | Confirmed |
| IMD | 9.74 | 9.68 | 8.28 | 11.89 | 1.00 | Confirmed |
| Education | 4.97 | 5.07 | 2.87 | 6.38 | 0.95 | Confirmed |
| Ethnicity | 4.83 | 4.90 | 3.08 | 6.73 | 0.86 | Confirmed |
| Age | 12.68 | 12.98 | 10.26 | 14.27 | 1.00 | Confirmed |
| Gender | 11.79 | 11.70 | 10.17 | 13.61 | 1.00 | Confirmed |
| Restaurants**** | 20.09 | 19.86 | 18.99 | 21.83 | 1.00 | Confirmed |
| Pubs | 6.49 | 6.39 | 4.83 | 7.73 | 1.00 | Confirmed |
| Fast food | 12.58 | 12.33 | 11.51 | 14.85 | 1.00 | Confirmed |
| Entertainment | 7.80 | 8.01 | 5.65 | 9.05 | 1.00 | Confirmed |
| Items > 1345 | 22.06 | 21.90 | 22.01 | 23.43 | 1.00 | Confirmed |
| Drinks > 100 | 17.70 | 17.76 | 14.95 | 20.01 | 1.00 | Confirmed |
| Items >600 | 24.07 | 24.14 | 21.25 | 26.02 | 1.00 | Confirmed |
| Number of mentions of milk | 11.42 | 11.59 | 9.95 | 12.48 | 1.00 | Confirmed |
| Number of mentions of salad | 16.18 | 16.44 | 14.19 | 17.96 | 1.00 | Confirmed |
| Number of unique vegetables | 17.40 | 17.38 | 14.55 | 19.09 | 1.00 | Confirmed |
| Number of desserts | 16.90 | 16.98 | 14.01 | 20.26 | 1.00 | Confirmed |

*weekend vs weekday
**Time of day customers visited the restaurant
***2021 vs 2022
****New variables were created for restaurants (vs. all other outlet types) and fast food (vs all other outlet types) and so on.

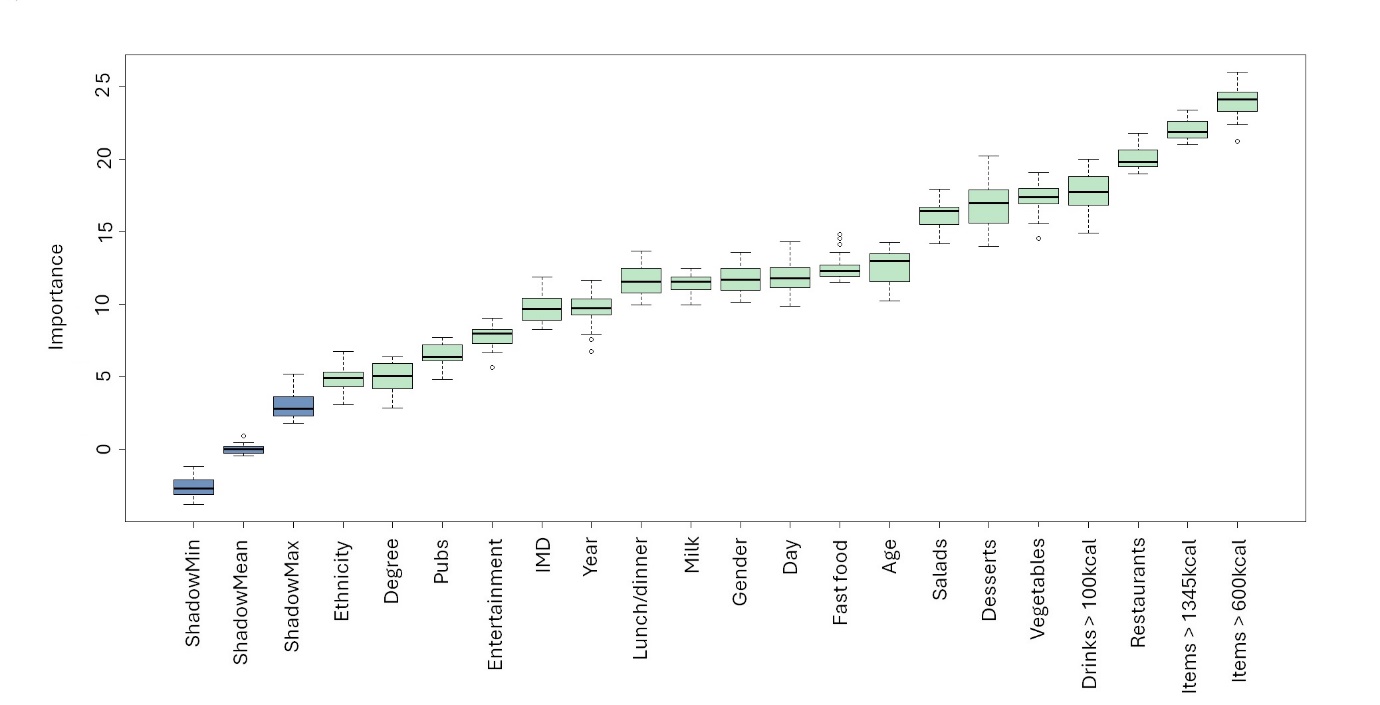
**Supplementary Material 4: Random Forest Model Feature Importance – with collinear variables removed**

**Supplementary Material 5: Interactions between menu healthiness scores with education and outlet type on energy consumed**

*Supplementary Table 3: Interactions between menu healthiness scores and education*

|  | **Kcal consumed** | | |
| --- | --- | --- | --- |
| *Predictors** | *Estimates* | *CI* | *p* |
| (Intercept) | 838.53 | 678.17 – 998.88 | **<0.001** |
| Menu healthiness score | -30.02 | -50.94 – -9.10 | **0.005** |
| Menu healthiness score × education | 2.93 | -17.60 – 23.45 | 0.780 |
| Observations | 3718 | | |
| R^2^ / R^2^ adjusted | 0.326 / 0.323 | | |
| (Intercept) | 694.91 | 459.13 – 930.69 | **<0.001** |
| Deep learning score | -8.67 | -40.32 – 22.98 | 0.591 |
| Deep learning score × education | 6.00 | -30.97 – 42.97 | 0.750 |
| Observations | 3718 | | |
| R^2^ / R^2^ adjusted | 0.316 / 0.313 | | |

*Models controlled for participant characteristics (age, gender, ethnicity, education) and situational characteristics (year, time of day, weekday/weekend, outlet IMD, outlet type).

*Supplementary Table 4: Interactions between menu healthiness scores and outlet type*

|  | **Kcal consumed** | | |
| --- | --- | --- | --- |
| *Predictors** | *Estimates* | *CI* | *p* |
| (Intercept) | 640.54 | 530.91 – 750.18 | **<0.001** |
| Menu healthiness score | -1.96 | -13.91 – 9.99 | 0.748 |
| **Outlet type Entertainment | -333.40 | -554.50 – -112.31 | **0.003** |
| **Outlet type Fast Food | 1406.07 | 1029.25 – 1782.89 | **<0.001** |
| **Outlet type Pubs, bars and inns | -523.48 | -701.29 – -345.67 | **<0.001** |
| **Outlet type Restaurant | 826.59 | 663.37 – 989.81 | **<0.001** |
| scores × outlet type Entertainment | 71.63 | 6.96 – 136.30 | 0.030 |
| scores × outlet type Fast Food | -187.77 | -243.16 – -132.38 | **<0.001** |
| scores × outlet type Pubs, bars and inns | 282.71 | 243.47 – 321.94 | **<0.001** |
| scores × outlet type Restaurant | -25.49 | -49.78 – -1.20 | 0.040 |
| Observations | 3718 | | |
| R^2^ / R^2^ adjusted | 0.357 / 0.353 | | |
| *Predictors* | *Estimates* | *CI* | *p* |
| (Intercept) | 486.01 | 178.07 – 793.94 | **0.002** |
| Deep learning score | 21.26 | -20.43 – 62.95 | 0.318 |
| **Outlet type Entertainment | 4145.95 | 723.11 – 7568.78 | 0.018 |
| **Outlet type Fast Food | 558.04 | 205.64 – 910.44 | **0.002** |
| **Outlet type Pubs, bars and inns | -1580.82 | -2054.61 – -1107.03 | **<0.001** |
| **Outlet type Restaurant | 489.43 | -296.29 – 1275.16 | 0.222 |
| scores × outlet type Entertainment | -610.06 | -1107.73 – -112.39 | 0.016 |
| scores × outlet type Fast Food | -55.22 | -109.38 – -1.06 | 0.046 |
| scores × outlet type Pubs, bars and inns | 311.81 | 244.58 – 379.05 | **<0.001** |
| scores × outlet type Restaurant | 22.83 | -91.81 – 137.47 | 0.696 |
| Observations | 3718 | | |
| R^2^ / R^2^ adjusted | 0.324 / 0.320 | | |

*This model controlled for participant characteristics (age, gender, ethnicity, education) and situational characteristics (year, time of day, weekday/weekend, outlet IMD).
**Reference value for outlet type is Cafes and coffee shops

**Supplementary Material 6: Exploration of relationships between healthiness scores and outlet type on kcal consumed**

Models were run separately among participants visiting each of the five outlet types.

|  | Cafes and coffee shops  (N=1217) | Entertainment (N=121) | Fast Food (N=1572) | Pubs, bars and inns (N=20) | Restaurant  (N=788) |
| --- | --- | --- | --- | --- | --- |
| Deep  learning score | 17.55 (p=.334) 95% CI: 18.08 - 53.18 | -596.06 (p=.110) 95% CI: 1330.15 - 138.03 | -32.71 (p=.073) 95% CI: 68.54 - 3.11 | - | 31.87 (p=.488) 95% CI: 58.29 - 122.03 |
| Menu healthiness score | -2.68 (p=.675) 95% CI: -15.19 - 9.83 | 110.24 (p=.260) 95% CI: -82.82 - 303.30 | **-188.71 (p<.001) 95% CI: -247.11 - -130.31** | - | **-29.47 (p=.009) 95% CI: -51.60 - -7.34** |

*Models controlled for participant characteristics (age, gender, ethnicity, education) and situational characteristics (year, time of day, weekday/weekend, outlet IMD).
**Estimates are in kcal consumed
- There were not sufficient data from Pubs bars and inns to estimate this model

**Supplementary Material 7: Two-step linear regression; Step one with menu characteristics only**

A linear regression model with all characteristics deemed important in the absence of multi-collinearity identified that menu characteristics alone explained 28.9% of variance. Without controlling for participant and situational characteristics, the number of unique vegetables, the percent of items over 600kcal and 1345kcal were significant predictors of kcal consumed.

|  | **Kcal consumed** | | |
| --- | --- | --- | --- |
| *Predictors* | *Estimates* | *CI* | *p* |
| (Intercept) | 597.43 | 547.07 – 647.80 | **<0.001** |
| Number of desserts | -0.13 | -1.19 – 0.93 | 0.811 |
| Number of unique vegetables | 5.60 | 3.15 – 8.05 | **<0.001** |
| Number of mentions of salad | -1.38 | -3.62 – 0.86 | 0.228 |
| Number of mentions of milk | -9.60 | -24.49 – 5.29 | 0.206 |
| Percent items > 600 | 10.02 | 8.83 – 11.22 | **<0.001** |
| Percent drinks > 100 | -0.85 | -1.57 – -0.14 | 0.020 |
| Percent items > 1345 | 35.88 | 25.47 – 46.30 | **<0.001** |
| Observations | 3718 | | |
| R^2^ / R^2^ adjusted | 0.290 / 0.289 | | |
